# Impact of baseline cases of cough and fever on UK COVID-19 diagnostic testing rates: estimates from the Bug Watch community cohort study

**DOI:** 10.1101/2020.09.03.20187377

**Authors:** Max T Eyre, Rachel Burns, Victoria Kirkby, Catherine Smith, Spiros Denaxas, Vincent Nguyen, Andrew Hayward, Laura Shallcross, Ellen Fragaszy, Robert W Aldridge

## Abstract

**Background:** Diagnostic testing forms a major part of the UK’s response to the current COVID-19 pandemic with tests offered to people with a continuous cough, high temperature or anosmia. Testing capacity must be sufficient during the winter respiratory season when levels of cough and fever are high due to non-COVID-19 causes. This study aims to make predictions about the contribution of baseline cough or fever to future testing demand in the UK.

**Methods:** In this analysis of the Bug Watch prospective community cohort study, we estimated the incidence of cough or fever in England in 2018–2019. We then estimated the COVID-19 diagnostic testing rates required in the UK for baseline cough or fever cases for the period July 2020-June 2021. This was explored for different rates of the population requesting tests and four second wave scenarios and then compared to current national capacity.

**Results:** The baseline incidence of cough or fever in the UK is expected to rise rapidly from 154,554 (95%CI 103,083 – 231,725) cases per day in August 2020 to 250,708 (95%CI 181,095 – 347,080) in September, peaking at 444,660 (95%CI 353,084 – 559,988) in December. If 80% of baseline cough or fever cases request tests, average daily UK testing demand would exceed current capacity for five consecutive months (October 2020 to February 2021), with a peak demand of 147,240 (95%CI 73,978 – 239,502) tests per day above capacity in December 2020.

**Conclusions:** Our results show that current national COVID-19 testing capacity is likely to be exceeded by demand due to baseline cough and fever alone. This study highlights that the UK’s response to the COVID-19 pandemic must ensure that a high proportion of people with symptoms request tests, and that testing capacity is immediately scaled up to meet this high predicted demand.

## Introduction

In response to the spread of novel coronavirus SARS-CoV-2, the United Kingdom (UK) has implemented large-scale public health measures that aim to reduce transmission and contact rates in the population. Maintaining control through diagnostic testing and self-isolation will increasingly depend upon self-diagnosis based on an individual’s symptoms. Current National Health Service (NHS) guidance is that any person in the community who develops at least one symptom of: a new continuous cough, a high temperature, or a loss of, or change in, normal sense of taste or smell (anosmia), should schedule a swab test with NHS services for home delivery or visit a testing site and self-isolate for up to 10 days after onset of symptoms or until a negative test result is received [1–3]. The NHS Test and Trace service will trace and notify close recent contacts of anyone who tests positive for coronavirus to self-isolate for 14 days.

The importance of COVID-19 testing in the UK’s response to the current pandemic is apparent in the five-pillar testing strategy, described by the UK Government in April 2020. The first two pillars use swab-based testing and molecular diagnosis of COVID-19 using real-time PCR [4]. Pillar 1 of the testing strategy relates to the swab testing for health and care workers and those with a clinical need, carried out by Public Health England (PHE) and NHS labs. Pillar 2 concerns swab testing for the wider population including social care and is carried out with commercial partners. The UK Government stated that an important marker for easing control measures and restrictions included having confidence that operational challenges, such as testing capacity, were “*in hand, with supply able to meet future demand*” [5].

Fever and cough are common symptoms in other acute respiratory viruses [6]. As a result of the non-specific nature of these respiratory symptoms, a large number of individuals meeting the UK’s COVID-19 diagnostic testing criteria – and being subsequently tested – will have cough and/or fever caused by a non-COVID-19 infection. It is therefore important to estimate the total number of cases in the population that would meet the diagnostic testing criteria (including both COVID-19 and non-COVID-19 cases) and the proportion of these cases that would seek testing, in order to ensure sufficient diagnostic testing capacity.

Bug Watch was a prospective community cohort study conducted in England in 2018–2019 that collected daily information on symptoms of a range of acute common infections [7]. Data collected within the study allows us to quantify the community incidence of fever and cough symptoms in England and describe seasonal patterns across a calendar year. Our study has two main objectives: first, to use Bug Watch data to estimate the all-age monthly incidence of cough or fever in England in the period 2018–2019; second, to estimate the UK COVID-19 diagnostic testing demand under current government testing policy for July 2020 – June 2021.

## Methods

### Study design, recruitment and data collection

Bug Watch was an online prospective community cohort study in England. Full details of the study design, recruitment and data collection are described in the protocol [7]. In brief, participants were recruited through an invitation letter sent to adults who participated in the 2013, 2014 and 2015 Health Survey for England (HSE). Parents or guardians were asked to register their children under 16 and complete surveys on their behalf. Any other adults within the same household were invited to register separately. Recruitment was conducted in four waves in March, June, September and November 2018. Data collected consisted of an online consent form and a baseline survey followed by weekly surveys sent by email to be completed by each participant. Each week, participants were asked to prospectively keep track of a wide range of symptoms of infection using a symptom diary. The primary outcomes of interest for this study were cough (defined as either a dry cough or coughing up phlegm) and fever. Each individual was followed up for six months. Only individuals with a 75% completion rate were included in the analysis.

Out of 19,741 adults who were invited to join the study, a total of 873 participants were included in the analysis (782 adults and 91 children that they had registered), providing a total follow-up time of 23,111 person-weeks. Cohort recruitment and baseline characteristics have been described in more detail [8], and are included in the supplementary material (S1).

### Ethics

Data were collected using Research Electronic Data Capture (REDCap)17 surveys hosted on the UCL Data Safe Haven, which is certified to the ISO27001 information security standard and conforms to NHS Digital’s Information Governance Toolkit. This study was given ethical approval by the UCL Research Ethics Committee (ID 11813/001).

### Statistical analysis

#### Baseline incidence of cough or fever in England

The first ten days of follow-up were excluded for all participants to remove prevalent infection syndromes. For participants reporting cough or fever symptoms within these first ten days, followup was started on the first day after this period with no symptoms. Incident cough or fever were defined as i) when a participant reported cough or fever for the first time; or ii) when either symptom was reported after a period of at least 10 days without symptoms. Non-specific symptoms could extend the duration of a cough or fever infection period.

Monthly adjusted incidence rates per 100,000-person-week for cough or fever were calculated for England, weighting to the mid-2019 population structure of England for age, sex and region [9] by post-stratification (using the R ‘survey’ package [10]). Monthly age-specific incidence rates per 100,000-person-week for cough or fever in England were calculated, weighting by sex and region, and are included in the supplementary material (S2).

#### UK testing demand due to baseline cough and fever cases

Monthly all-age adjusted incidence rates of baseline (non-COVID-19) cough or fever were estimated for the UK, weighting to the mid-2019 population structure of the UK for age and sex [9]. These rates were used to estimate the average number of individuals in the UK with an incident case of non-COVID-19 cough or fever each day for each month in the period July 2020 – June 2021.

Predictions for the daily testing demand expected in the UK between July 2020 and June 2021 due to baseline cough or fever cases were made based on our incidence estimates. We assumed that individuals only request a test on the first day that they experience symptoms. We explored a range of scenarios for the proportion of cough or fever cases that request a test (PROPTEST). Four values were explored: 40%, 60%, 80% and 100%. The predicted impact of baseline cough or fever cases on UK testing capacity was calculated as the difference between current UK Pillar 1 and 2 laboratory testing capacity estimates and predicted testing demand between July 2020 and June 2021 based on these scenarios. Capacity estimates used in this analysis were reported by the UK government for the period 6–12th August 2020 as 1,459,418 tests per week for Pillars 1 and 2, and 880,000 tests per week for only Pillar 2 [11].

#### Total UK testing demand including symptomatic COVID-19 cases

Four scenarios (C1-C4) for additional demand due to a second COVID-19 wave in the UK during winter 2020–2021 were explored. A range of average daily incidences for COVID-19 cases for each month between July 2020 and June 2021 were considered to reflect uncertainty about future COVID-19 transmission levels, from the lowest in scenario C1 to the highest incidences in scenario C4. These scenarios follow a similar epidemic curve shape to predictions reported in the Academy of Medical Sciences’ report “Preparing for a Challenging Winter 2020/21”, with the highest incidences in January and February 2021 and the peak incidence in our worst-case scenario (C4) equal to the peak incidence predicted in the report for a reproductive number R_t_ = 1.5 between September 2020 to July 2021. More information about exact derivation of the incidences used in these scenarios can be found in the supplementary material (S5). Based on estimates from the COVID Symptom Study app, 87.5% of these cases of COVID-19 were assumed to exhibit symptoms of cough, fever or loss of smell or taste [12] with the proportion of these symptomatic cases expected to request tests explored using the PROPTEST parameter. Total demand for swab tests due to baseline cough and fever cases and COVID-19 illnesses was calculated as:

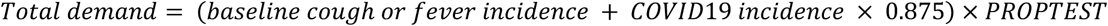

The total predicted testing demand in each month was then calculated for each COVID-19 scenario (C1-C4).

All analyses were conducted using R [13].

## Results

### Cough or fever incidence in England

Out of a total of 585 episodes of cough or fever, participants experienced 431 (73.7%; 431/585) episodes of cough, 57 (9.7%; 57/585) of fever and 97 (16.6%; 97/585) episodes with both cough and fever symptoms.

Monthly age-, sex- and region-adjusted incidence rates of cough or fever per 100,000-person-week and 95% confidence intervals are shown for the 12-month study period in England in Figure 1. There was clear seasonal variation in incidence, with the lowest rates in June and highest rates in December with 1,333 (95%CI 753 – 2,361) and 4,958 (95%CI 3,847 – 6,390) incident episodes of cough or fever per 100,000-person-week, respectively. The high incidence in December coincides with UK public holidays and lower temperatures, when indoor contact rates are higher.

**Figure 1.**
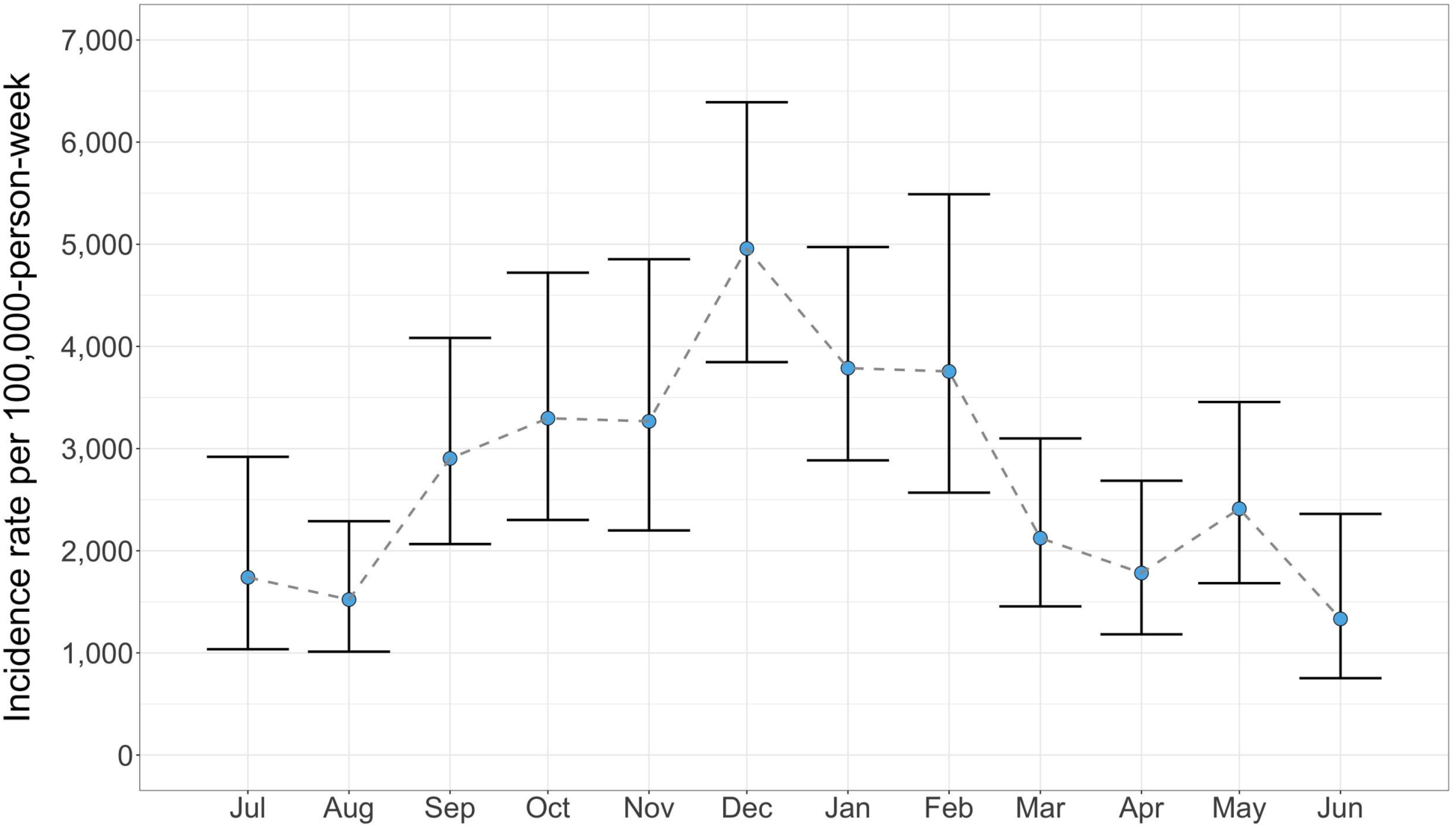
Monthly adjusted incidence rates of cough or fever per 100,000-person-week in England with 95% confidence intervals. Weighted to the mid-2019 population structure of England by age, sex and region.

### UK testing demand due to baseline cough and fever cases

Predictions for the average daily number of baseline (non-COVID-19) cough or fever cases in the UK between July 2020 – June 2021 are shown for each month in Figure 2. Under current UK government policy, all of these cases would be entitled to a COVID-19 swab test. After the lower incidence summer period of 2020, the incidence starts to rise rapidly, increasing from 154,554 (95%CI 103,083 – 231,725) cases per day in August to 250,708 (95%CI 181,095 – 347,080) in September, before peaking at 444,660 (95%CI 353,084 – 559,988) daily cases in December. This high incidence continues to exceed current UK laboratory testing capacity for Pillars 1 and 2 until the end of winter before falling to 204,750 (95%CI 141,392 – 296,499) cases per day in March 2021.

**Figure 2.**
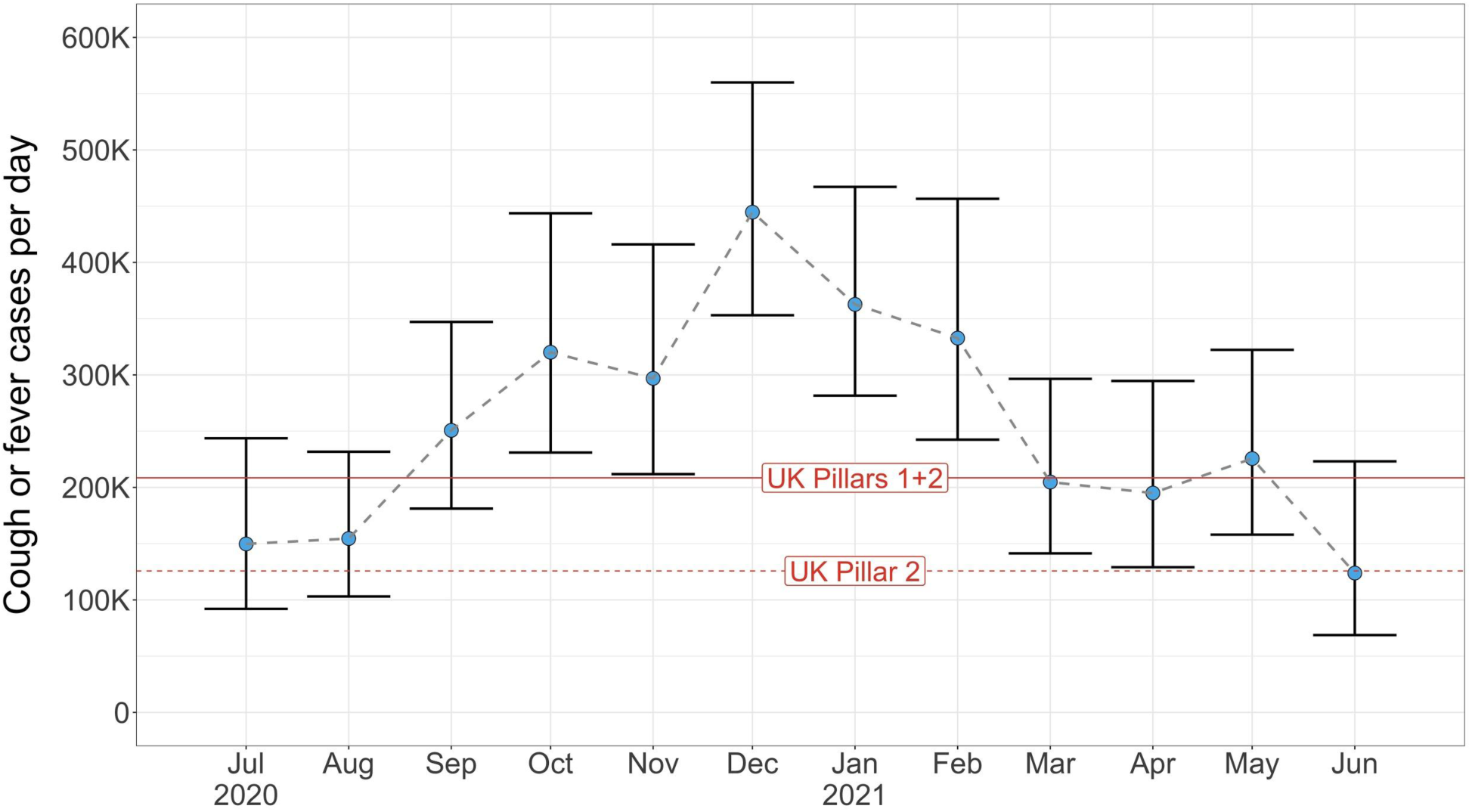
Predicted baseline number of individuals (in thousands) in the UK with an incident case of cough or fever each day shown for each month. Current laboratory capacity (measured as daily tests) for Pillars 1 and 2 and Pillar 2 in the UK are shown as solid and dashed red lines, respectively.

Remaining UK Pillar 1 and 2 capacity after testing baseline cough or fever cases is shown in Figure 3 for four values of the proportion of cough or fever cases which request tests (PROPTEST). The peak in cases in the autumn and winter of 2020–2021 is likely to place significant stress on the UK’s testing service in these months. Figure 3a shows that when only 40% of these cases request tests, current capacity is sufficient for the entire year. However, as this proportion increases, current capacity becomes insufficient for predicted demand. When 60% request tests, demand in December 2020 and January 2021 exceeds current capacity by 58,308 (95%CI 3,362 – 127,505) and 9,121 (95%CI –39,541 – 71,798) tests per day (Figure 3b). For 80% (Figure 3c), the daily average demand for tests exceeds current capacity in five consecutive months (October 2020 to February 2021), with a peak of 147,240 (95%CI 73,978 – 239,502) tests per day above capacity expected in December 2020. If all individuals experiencing non-COVID-19 cough or fever request a test, we estimate that there will only be a significant capacity surplus in the summer months of July and August 2020 and June 2021.

**Figure 3.**
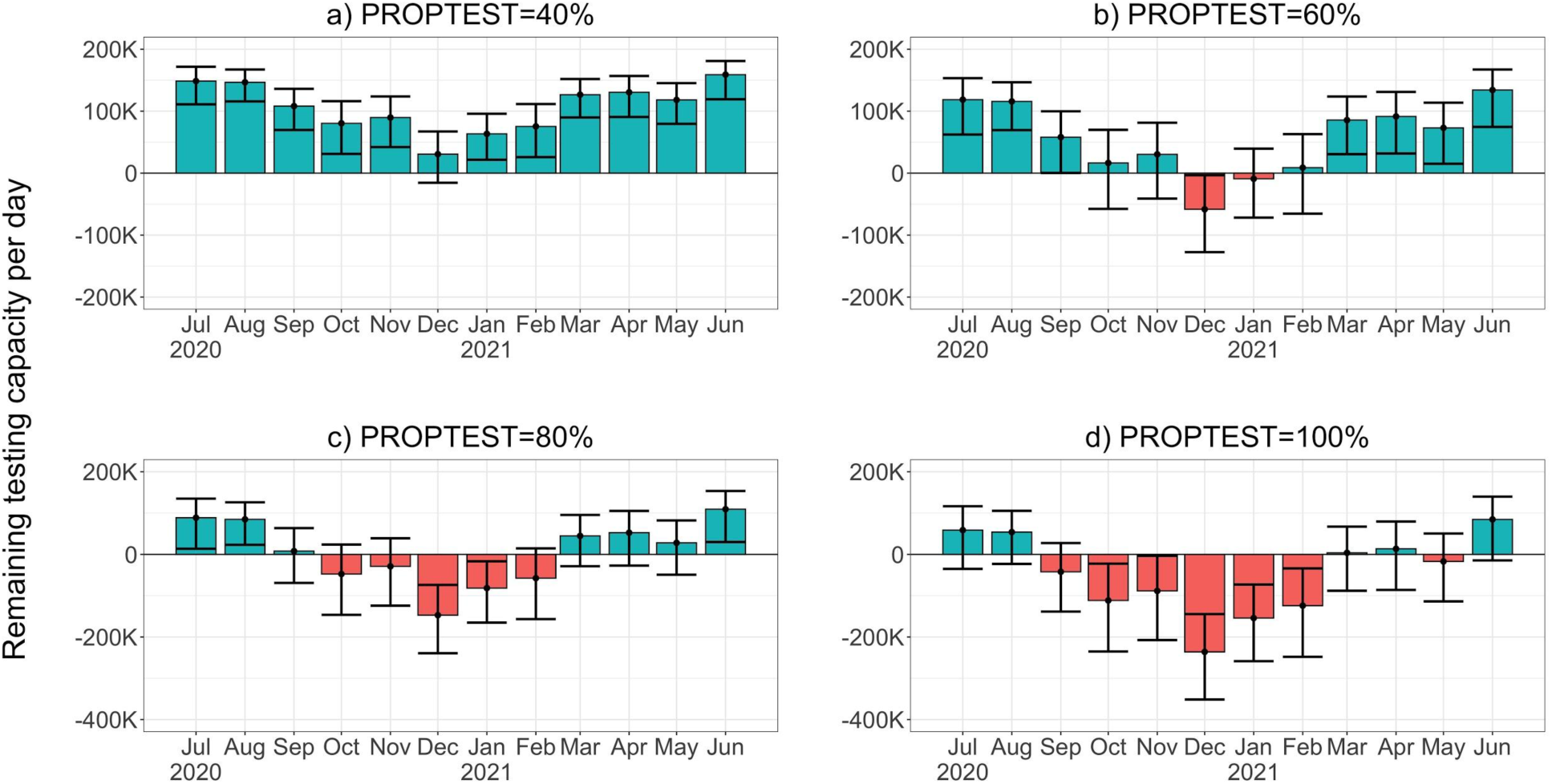
Remaining UK Pillar 1 and 2 testing capacity (thousands of tests) available after testing baseline (non-COVID-19) cases of cough or fever in the UK each day. Panels a) to d) show results for four values of the proportion of cough of fever cases requesting tests (PROPTEST). Blue and red bars indicate that cough or fever testing demand is within or in excess of current capacity, respectively.

### Total UK testing demand including symptomatic COVID-19 cases

The four scenarios (C1-C4) for a winter COVID-19 epidemic in the UK which were explored in this study are shown in Figure 4. All scenarios follow the same epidemic curve shape with the highest average daily incidences between December 2020 and March 2021 and peak incidences in January of 18,134 (in scenario C1), 36,268 (C2), 54,402 (C3) and 72,536 (C4) cases per day.

**Figure 4.**
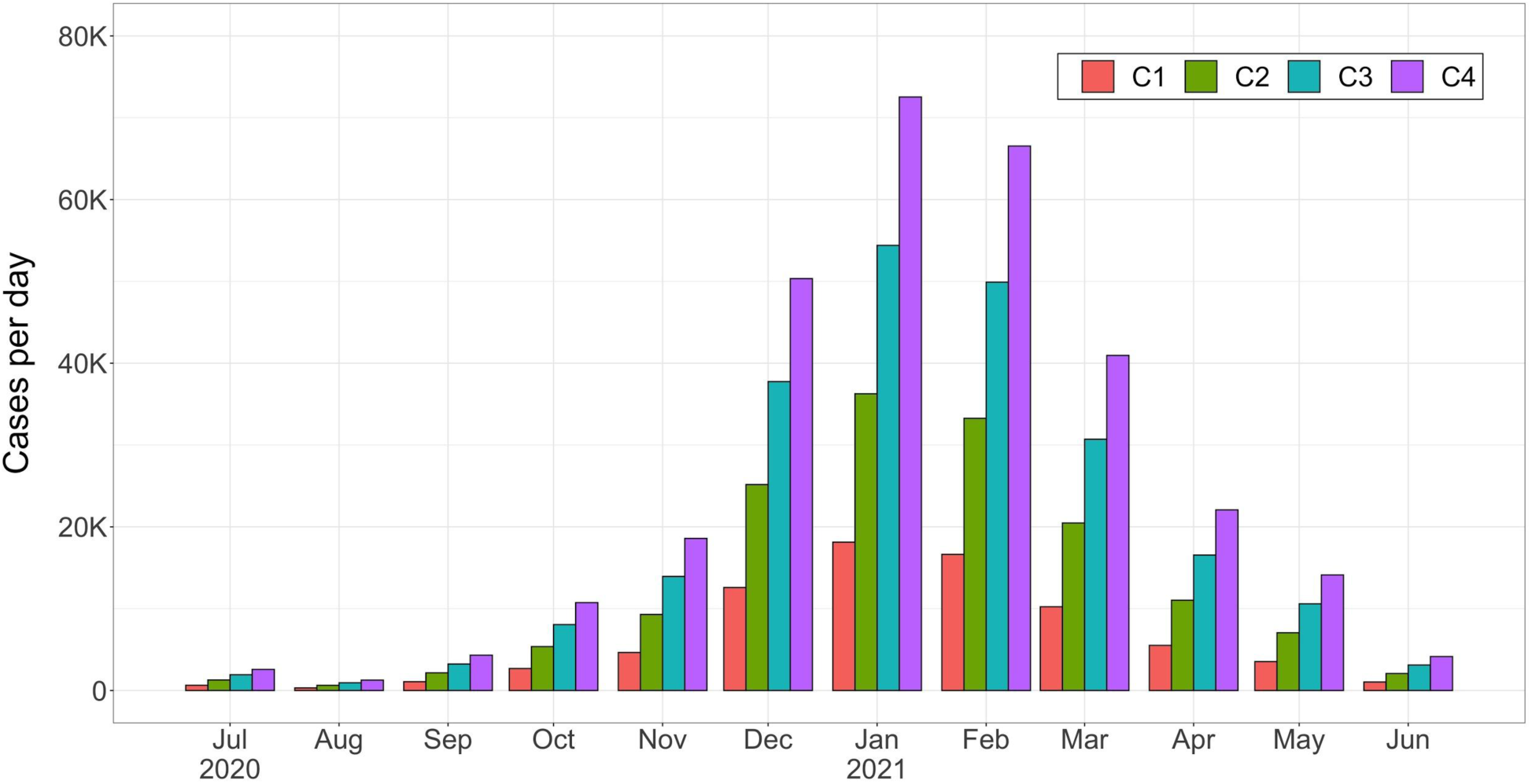
Four scenarios (C1 to C4) for average daily COVID-19 incidence in the UK shown for each month

For the midrange C2 scenario, the remaining testing capacity available after testing baseline cough or fever cases and symptomatic COVID-19 cases is shown in Figure 5 (results for all other scenarios were similar and are included in the supplementary material S3). These results were similar to those for baseline cough or fever cases only (Figure 3). Capacity is not predicted to be exceeded when only 40% of cough or fever cases and symptomatic COVID-19 cases request tests. When 60% of cases request tests, there is a predicted daily demand in December 2020 of 71,522 (95%CI 16,576 – 140,719) tests above capacity and the additional COVID-19 demand pushes total demand in February 2021 above capacity. When 80% of cases request tests, we see an increased deficit during the epidemic’s peak months of December to January and, when 100% request a test, UK testing capacity is predicted to be severely strained from September 2020 to May 2021.

**Figure 5.**
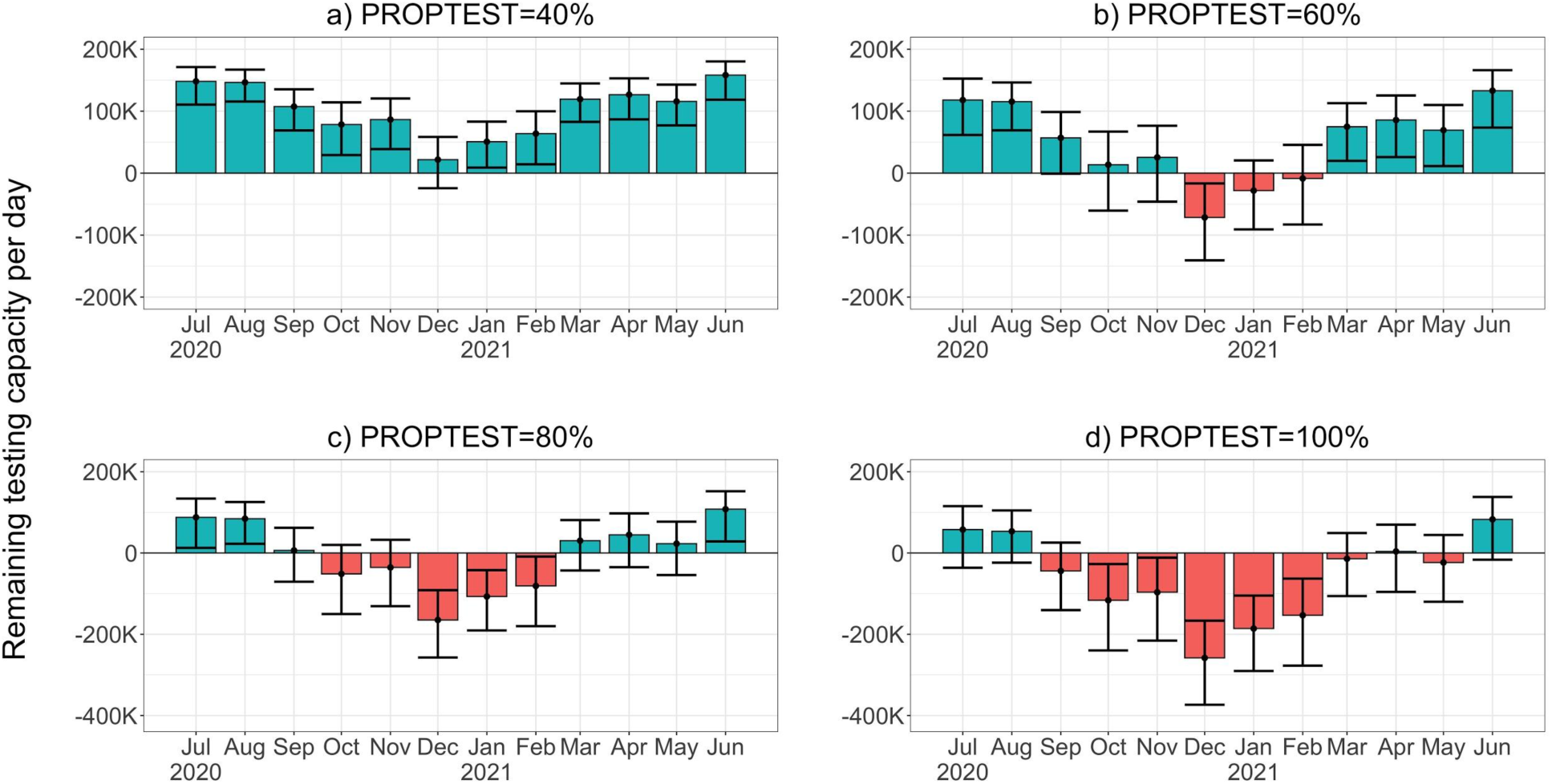
Remaining UK Pillar 1 and 2 testing capacity (thousands of tests) per day after testing baseline (non-COVID-19) cases of cough or fever and symptomatic COVID-19 cases in the UK each day for scenario C2. Panels a) to d) show results for four values of the proportion of cases requesting tests (PROPTEST). Blue and red bars indicate that testing demand is within or in excess of current capacity, respectively. Note that panel a) has a different y-axis scale to panels b) – d).

The relatively small contribution of symptomatic COVID-19 cases to total predicted testing demand in the UK for scenario C2 was also found for all other scenarios, shown in Figure 6, for an assumed value of 80% of cases requesting a test. The more severe C3 and C4 scenarios result in higher total demand in January 2021 of 328,227 (95%CI 263,345 – 411,797) and 340,921 (95%CI 276,039 – 424,491) tests per day, respectively, relative to 302,839 (95%CI 237,957 – 386,409) in the C1 scenario. While these increases are not negligible, the total testing demand is predominantly driven by baseline cough or fever cases, and the overall effect of increased COVID-19 transmission between these scenarios is small. For example, in the highest transmission scenario, C4, and peak COVID-19 incidence in January 2021, only 14.9% of tests are expected to be requested by COVID-19 cases. This is a result of the high incidence of baseline cough or fever cases during the second wave peak months of December 2020 to February 2021. Consequently, relative to current UK testing capacity, the overall picture remains the same across the next year for all four scenarios, with the period October to February at high risk of surpassing current UK testing capacity. As the proportion of cases requesting tests is applied to both baseline and symptomatic COVID-19 cases, symptomatic COVID-19 cases contribute the same proportion of the total testing demand for all other values of the proportions of cases requesting tests, although total demand varies significantly with this proportion (see supplementary material S4).

**Figure 6.**
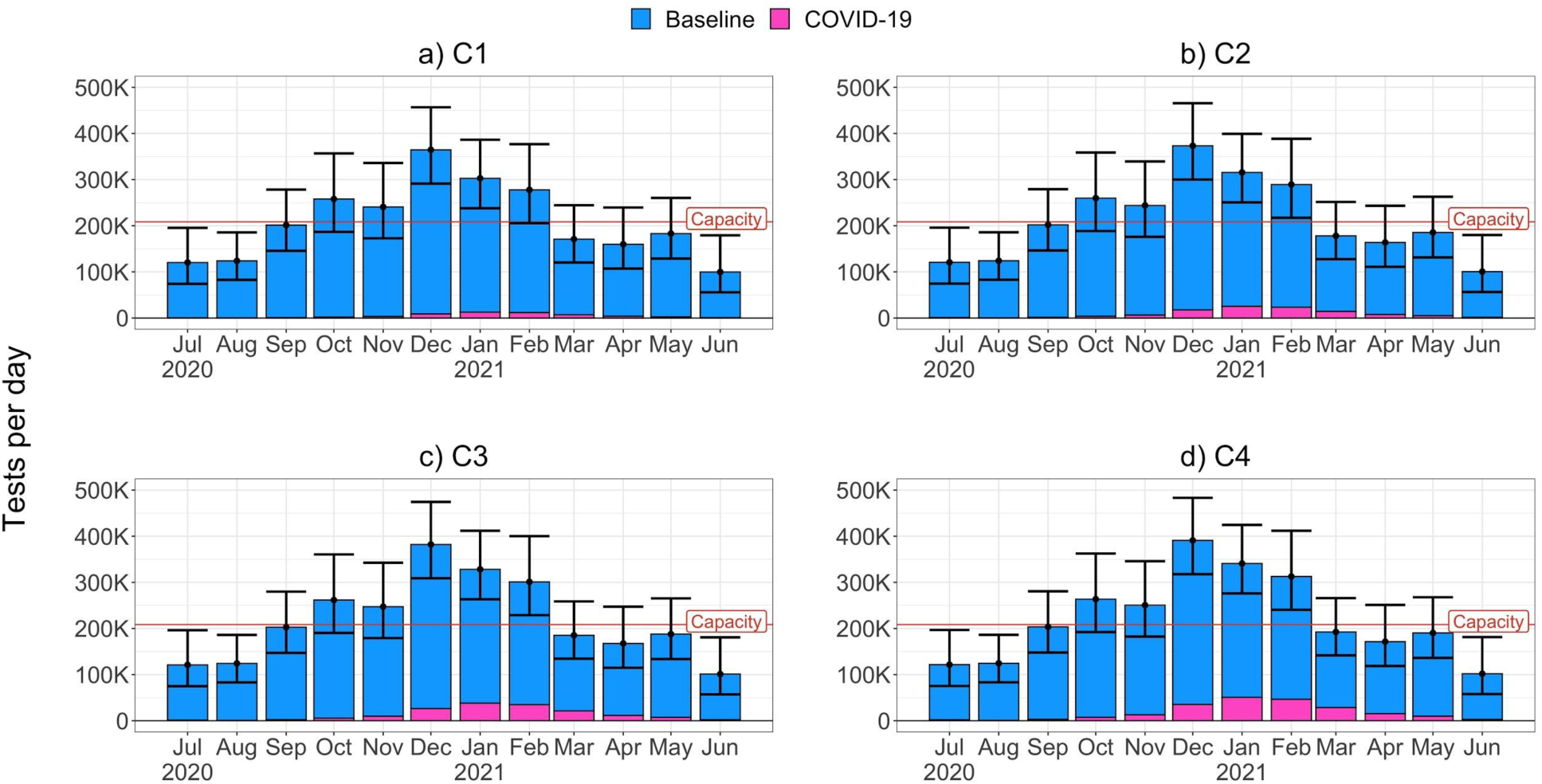
Predicted daily testing demand (thousands of tests) due to baseline cough or fever cases (blue) and symptomatic COVID-19 cases (pink) in the UK, assuming that 80% of cases request tests. Panels a) to d) show results for scenarios C1 to C4. Current daily UK Pillar 1 and 2 testing capacity is marked with a red line.

## Discussion

In this study, we estimate the baseline incidence of cough or fever cases and their potential impact on COVID-19 diagnostic testing services in the UK in July 2020 – June 2021. Our results show that a high baseline incidence of cough and fever between October 2020 and February 2021 will place a significant strain on UK testing capacity. We estimate that if more than 80% of people with symptoms request a test, daily demand for COVID-19 swab tests will exceed current capacity by a significant margin for these five months. We find that testing demand will be predominantly driven by baseline cough or fever incidence, rather than symptomatic COVID-19 cases, and that the proportion of people with symptoms who request a test is a key determinant of demand. To our knowledge, this is the first paper published quantifying baseline cough or fever cases in the UK and their impact on COVID-19 diagnostic testing services.

The strong seasonal trend in baseline incidence of cough or fever cases in the UK is a clear indicator of the challenges which the UK’s testing services will face in the autumn and winter months of 2020–2021. Our results show that while the UK’s current diagnostic testing capacity may be sufficient for July to September 2020, this is unlikely to be the case from October 2020. If capacity is exceeded to the extent predicted in our results, a large backlog of unprocessed tests can be anticipated and a significant proportion of COVID-19 positive cases are likely to remain untested. Prompt identification of cases is critical for effective contact tracing and real-time visualisation of epidemiological trends to inform national and local-level public health interventions. It is therefore imperative that the UK’s testing capacity is scaled up immediately to ensure that there is sufficient capacity to respond to this predicted rise in testing demand and ensure that detection of COVID-19 cases is not compromised. Delays in testing, due to lack of capacity, may also disincentivise people from getting tested and may result in unnecessarily extended self-isolation of COVID-19 negative households. The UK testing strategy has acknowledged the need to expand testing capacity rapidly [14] and the Government has announced their ambition to increase antigen testing capacity to half a million tests per day by the end of October 2020 [15]. Our calculations show that if this additional capacity is achieved, it would significantly reduce the risk of testing backlogs over the next six months. However, it is unlikely to be sufficient to cover routine testing of asymptomatic health and care professionals which may be implemented if infection risk rises in September or October [16].

Our results show that testing demand is likely to be driven by the proportion of people with symptoms that request a test. An effective public health response to COVID-19 in the UK requires all individuals experiencing symptoms to request a test promptly after symptoms begin and our estimates for higher values of the proportion requesting tests should be considered as a necessary requirement for the UK’s testing capacity. Therefore, in addition to scaling up capacity, the UK’s response must also ensure that a high proportion of symptomatic people are requesting tests to begin with. The high baseline incidence of cough or fever cases reported in our analysis highlights the scale of this issue, and culturally and linguistically appropriate public engagement campaigns, as well as accessible and rapid test-ordering systems, will be critical to a successful response.

In this study, we assume that the baseline incidence of cough or fever during the study period of 2018–19 is representative of 2020–2021. While easing of public health interventions has occurred in most of the UK during July-August 2020, many measures, such as social distancing and bans on mass gatherings, will continue. Conceivably, this may already be impacting upon the incidence of other respiratory pathogens, with UK surveillance data from 2019 to 2020 suggesting low levels of influenza activity in the community [17]. However, as the World Health Organization has noted [18], these early results should be interpreted with caution, as the latter part of this influenza season coincided with the increased incidence of COVID-19. This has resulted in several respiratory surveillance indicators being affected – such as GP influenza-like illness consultation rates – owing to changes in health-seeking behaviour and reporting practices. Additionally, given that children and young adults are thought to drive transmission of influenza and other seasonal respiratory infections, the reopening of schools and universities in the UK in September may maintain baseline cough or fever incidence at 2018–19 levels. It is consequently difficult to determine whether our calculations overestimate the future incidence of baseline cough or fever cases with an infectious aetiology. However, our analysis of lower proportions of the population requesting tests shows that lower levels of baseline cough or fever incidence, relative to 2018–2019 levels, would still place a significant strain on current testing capacity.

The COVID-19 transmission scenarios explored in this study are speculative and reflect uncertainty about the potential size and timing of a second COVID-19 wave. However, they present a range of reasonable scenarios based on previous modelling predictions [19] that are consistent with the expectation that a second wave will have a lower peak incidence and flatter epidemic curve than the first wave in March – July 2020 in the UK [20]. Our results show that total testing demand is relatively insensitive to COVID-19 transmission and that, even in more severe scenarios, testing demand due to baseline cough or fever cases will outweigh demand due to symptomatic COVID-19 cases.

A limitation of our study was that participants were more likely to be older, female, healthier and living in less deprived areas than the general population of England. To account for age and sex, we adjusted for these variables in our incidence estimates. There were also a disproportionately low number of Black, Asian and other Minority Ethnic (BAME) participants included in the study – communities that are known to have been particularly adversely affected by COVID-19 [21]. Consequently, our results do not account for possible differences in the incidence of baseline cough and fever in these underrepresented groups. Another possible limitation of this study is that a cough was defined as ‘incident’ rather than ‘continuous’ (lasting more than one hour or three or more coughing episodes in 24 hours), therefore potentially differing from the UK’s COVID-19 diagnostic symptomatology. We may consequently overestimate the number of cough cases who would require a test under current NHS guidance. We also note that our estimates of testing demand were largely driven by cough symptoms, as fever was comparatively less common. Data was not collected on altered or lost sense of smell or taste, but we expect this to be rare in comparison to symptoms of cough or fever.

In conclusion, our study provides estimates of the baseline incidence of cough or fever in the general population in the UK. Our calculations indicate that the UK’s current COVID-19 testing capacity is insufficient for high predicted demand between October 2020 and February 2021, even when only accounting for baseline cough or fever cases. It highlights the need to ensure that a high proportion of people with symptoms request tests and for testing capacity to be immediately scaled up – the Government’s ambition to increase antigen testing capacity to 500,000 tests per day must be a priority. Otherwise, compounded by high COVID-19 levels projected in a second wave, UK testing capacity could be overwhelmed leading to failure of the NHS Test and Trace service and an inability to control the further spread of COVID-19.

## Data Availability

Upon publication, data and R script used for this analysis and production of figures will be made available on Dryad.

## Author Contributions

Conceived the project: RWA, AH

Literature search: RB, MTE, VK

Study design and data collection: EF, CS, AH

Data analysis: MTE, RB

Figures: MTE

Data interpretation: MTE, RB

Wrote the original draft of the manuscript: MTE, RB, VK

Comments on all manuscript: all authors

